# Comparative Analysis of Stress Responses in Medical Students Using Virtual Reality Versus Traditional 3D-Printed Mannequins for Pericardiocentesis Training

**DOI:** 10.1101/2024.08.22.24312406

**Authors:** Alberto Rubio-López, Rodrigo García Carmona, Laura Zarandieta Román, Alejandro Rubio Navas, Ángel González Pinto, Pablo Cardinal-Fernández

## Abstract

**Background:** As medical education evolves, innovative methods like virtual reality (VR) and 3D-printed mannequins are increasingly used to simulate high-stress medical scenarios realistically. This study investigates the effectiveness of VR and 3D-printed mannequins in replicating stress levels during pericardiocentesis training, comparing their impact on the emotional and physiological responses of learners.

**Methods:** We enrolled 108 final-year medical students who were randomized to train with both VR and 3D-printed mannequins. Heart rate variability (HRV) analysis was employed to assess stress responses. Additionally, a secondary analysis examined the influence of demographic factors, lifestyle, medication use, and academic stress on these responses.

**Results:** Both VR and traditional mannequin-based training methods proved equally effective in simulating the stress levels encountered in real medical procedures. Our findings indicate significant interactions between stress markers and demographic factors, which highlights the complex nature of stress responses in medical education and underscores the necessity for personalized training approaches.

**Conclusion:** The study validates the use of VR as a viable alternative to traditional mannequins, capable of simulating the technical skills and emotional pressures of medical procedures such as pericardiocentesis. Incorporating VR into medical training programs may enhance learning outcomes and accessibility, particularly in settings constrained by resources.

## INTRODUCTION

The field of medical education has been revolutionized by the adoption of simulation-based learning, which aligns with the “learning by doing” [1] philosophy. This approach enhances patient safety by allowing students to practice clinical procedures in safe environments, offering reproducible scenarios for consistent learning outcomes while accommodating various educational levels. Despite its widespread benefits, the application of simulation-based learning is often hampered by the high costs of advanced simulators and the steep learning curves for instructors, which can exacerbate educational disparities, particularly in economically constrained settings [2].

In response to these challenges, recent advancements in 3D printing and virtual reality (VR) technologies have begun to bridge the gap [3]. 3D printing provides a cost-effective means to create detailed medical models [4], thereby broadening the scope and flexibility of simulations. Similarly, VR offers immersive and realistic environments that enhance both sensory and emotional training experiences [5], making these tools accessible across different educational settings and platforms such as Oculus and HTC.

Despite these technological advances, a critical gap persists in evaluating the emotional impact of these simulations, especially their ability to evoke authentic stress responses during high-stakes procedures [6]. This study addresses this gap by focusing on pericardiocentesis—a procedure notorious for its complexity and high stress levels [7]. We aim to compare the efficacy of traditional 3D-printed mannequins and VR simulations in eliciting realistic stress responses as measured by heart rate variability (HRV), a validated stress biomarker. Through this comparison, we seek to ascertain which simulation method better prepares medical students for the real-world pressures of medical emergencies, particularly in resource-limited settings [8].

## METHODS

### Study Design

This validation study builds on our pilot research to compare two simulation methods for pericardiocentesis training: a 3D-printed mannequin and a virtual reality (VR) setup using the Unity game engine. The primary objective was to evaluate and contrast the ability of these methods to simulate realistic emotional and physiological responses encountered in actual medical procedures.

### Randomization and Participant Grouping

We enrolled 132 final-year medical students from the CEU University School of Medicine in Madrid. Before participation, students completed demographic and health behavior questionnaires. Participants were randomly assigned into two groups using a computer-generated sequence, with one group starting with VR training followed by the mannequin training, and the other group starting in reverse order. This counterbalanced design aimed to mitigate any order effects on learning outcomes and stress responses.

### Development and Validation of the Training Models

The 3D-printed mannequin was designed using Tinkercad and Adobe Fusion 360 to ensure anatomical precision. It was printed with MeshMixer software, Ultimaker Cura for slicing, and an Ender 3 printer. Concurrently, the VR model was developed in Unity to mirror the mannequin’s design, ensuring a consistent and immersive training experience. Both models were rigorously validated by clinical experts from HM Montepríncipe University Hospital in Madrid.

### Ethical considerations and recruitment

The study received approval from the HM Montepríncipe University Hospital Research Ethics Committee (Code: 18.12.1339.GHM) and spanned from November 7, 2023, to February 12, 2024. All participants provided written informed consent, and their data were anonymized to ensure confidentiality.

### Inclusion and Exclusion Criteria

All consenting final-year medical students were eligible, except those on cardiac medication, consuming excessive caffeine, or simpaticomimetics. Students with incomplete data or improperly recorded signals were excluded from the analysis. Participants received standardized training through an instructional video on pericardiocentesis before the simulations.

### Stress response measurement

To assess stress responses, students were equipped with three electrodes on the torso to continuously monitor heart rate using the Biosignal Plux system. This setup was established prior to the initiation of simulation training to ensure robust data collection on physiological stress indicators. The collected data were time-stamped, allowing for the separate analysis of each part of the corresponding procedure. Data acquisition, processing, and conversion into values for statistical analysis were conducted using OpenSignals digital signal processing software

Heart rate variability (HRV) analysis was employed to assess stress, as it serves as a sensitive marker for dysregulation in the Autonomic Nervous System (ANS). HRV is defined as the temporal variation in the intervals between consecutive heartbeats over a predefined period, providing a quantitative measure of autonomic nervous system activity that is critical under stressful conditions.

The parameters used in our HRV analysis include:

- **Frequency-domain parameters**:
  - Low Frequency (LF): Primarily reflects sympathetic activation but also includes parasympathetic influences. It is used to assess the balance of autonomic input.
  - High Frequency (HF): Acts as a specific indicator of parasympathetic nervous system (PNS) activity and is linked to stress relief mechanisms.
  - LF/HF Ratio: Serves as an indicator of the balance between sympathetic and parasympathetic activity, with higher values indicating increased stress levels.
- **Time-domain parameters**:
  - Root Mean Square of Successive Differences (rMSSD): Measures the short-term variability in heart rate and is closely related to the parasympathetic nervous system’s control over heart rate.
  - Percentage of Successive NN Intervals that Differ by More than twenty ms (PNN20) and fifty ms (PNN50): These parameters provide additional insight into the variability of heart rate, correlating strongly with parasympathetic activity.
- **Non-linear parameters**:
  - SD1/SD2 Ratio: Derived from the Poincaré plot, which visually represents the temporal intervals between consecutive heartbeats. This ratio correlates with the LF/HF ratio and provides insight into the complexity of heart rate variability patterns, reflecting stress levels.

These measures collectively provide a comprehensive profile of the students’ stress responses during simulation training, offering valuable insights into how different simulation modalities impact the autonomic nervous system under stress-inducing conditions

### Secondary Analysis of Stress Indicators

We conducted a secondary analysis to examine the effects of external factors such as medication intake, lifestyle habits, and academic pressures on stress responses. This analysis helped contextualize the physiological data, offering deeper insights into the multifactorial influences on stress levels during clinical training

### Data Collection Phases

- **Baseline Data Collection**: Initial stress data were collected in a resting state to establish baseline levels for each participant.
- **First Training Scenario**: Students underwent training with either the mannequin or VR, followed by a resting phase to mitigate carryover effects.
- **Second Training Scenario**: Participants then trained with the alternate method, facilitating a comparative analysis of stress responses

### Statistical analysis

Stress parameters between groups (VR-First and Mannequin-First) were compared using IBM SPSS software version 20. Analysis included:

- **Normality Testing**: Kolmogorov–Smirnov test to assess data distribution.
- **Comparative Analysis**: Friedman test for intra-group changes over conditions (baseline, simulation, rest, VR).
- **Paired Comparisons**: Friedman test and Wilcoxon signed-rank test for specific condition comparisons.
- **Secondary Analysis**: Repeated-measures ANOVA to explore relationships between stress responses and demographic factors, with post hoc analyses using Bonferroni and Tukey corrections.

## RESULTS

### Demographic and lifestyle characteristics

Our study initially enrolled 132 medical students, which after screening for eligibility criteria, was reduced to 108 participants. These students were equally divided into two groups of fifty-four. The cohort consisted of female students (87%) with an average age of 23.6 years. A significant portion of participants (78%) reported no prior experience with VR headsets. Comprehensive assessments of lifestyle factors—such as living arrangements, relationship status, income levels, familial responsibilities, and work-study balance—were conducted to understand the external influences on stress (Table 1). The impact of medication usage (e.g., anxiolytics, antidepressants) and lifestyle choices (smoking, alcohol consumption, caffeine intake, physical activity) were also examined (Table 2). Additionally, academic factors such as study hours, perceived stress levels, and mental health concerns were evaluated (Table 3)

**Table 1.** Lifestyle factors.

| **Characteristic** | **Frequency (%)** |
| --- | --- |
| **Marital Status** |  |
| Single | 50 |
| In a Relationship or married | 50 |
| **Living Arrangement** |  |
| With Parents or partner | 69.45 |
| Alone or shared apartment | 30.55 |
| **Family Income Level (€/Monthly)** |  |
| < 2000 to 3000 | 17.59 |
| 3000-5000 | 52.78 |
| > 5000 | 29.63 |
| **Working and Studying** |  |
| Yes | 22.22 |
| No | 77.78 |
| **Family Responsibilities** |  |
| Yes | 17.59 |
| No | 82.41 |

**Table 2.** Medication and Substance Use.

| **Characteristic** | **Frequency (%)** |
| --- | --- |
| **Smoking Habits** |  |
| Never Smoked | 77.78 |
| Smokes occasionally | 22.22 |
| **Regular Alcohol Consumption** |  |
| Never | 14.81 |
| Occasionally/Socially | 84.26 |
| Daily | 0.93 |
| **Coffee Consumption** |  |
| None or < 1 Cups/Day | 96.3 |
| >3 Cups/Day | 3.70 |
| **Physical Activity** |  |
| Never | 7.41 |
| Occasionally | 44.44 |
| Regularly | 48.15 |
| **Recreational Drug Use** |  |
| Yes | 0.93 |
| No | 99.07 |
| **Energy Drinks Consumption** |  |
| Yes | 1.85 |
| No | 98.15 |
| **Anxiolytic Medication** |  |
| Yes | 5.56 |
| No | 94.44 |
| **Antidepressant Medication** |  |
| Yes | 4.63 |
| No | 95.37 |

**Table 3.** Academic and mental health factors.

| **Characteristic** | **Frequency (%)** |
| --- | --- |
| **Daily Study Hours** |  |
| <1 to 2 | 8.76 |
| 3 to 5+ | 91.24 |
| **Belief that Stress Affects Academic Performance** |  |
| Yes | 75 |
| No | 25 |
| **Considered Dropping Out Due to Stress** |  |
| Yes | 41.67 |
| No | 58.33 |
| **Considered Self-Harm During Studies** |  |
| Yes | 14.81 |
| No | 85.19 |
| **Perceived Well-Being Level (0 Very Poor/10 Very Good)** |  |
| 0 to 5 | 8.33 |
| 6 to 8 | 72.2 |
| 9-10 | 19.44 |

### Biological stress analysis

Heart rate variability (HRV) analysis was employed to assess stress levels across four distinct states: basal, during VR simulation, resting, and during mannequin simulation. The Friedman test was used to analyze differences in biometric parameters across these states, revealing significant changes in HRV parameters (p < 0.01) from the baseline rest state to the active simulation states. This significant variation across time, frequency, and non-linear domains of HRV data underscores the efficacy of our biometric markers in detecting stress under different conditions, as detailed in Table 4 and Figures 1,2.

**Table 4.**
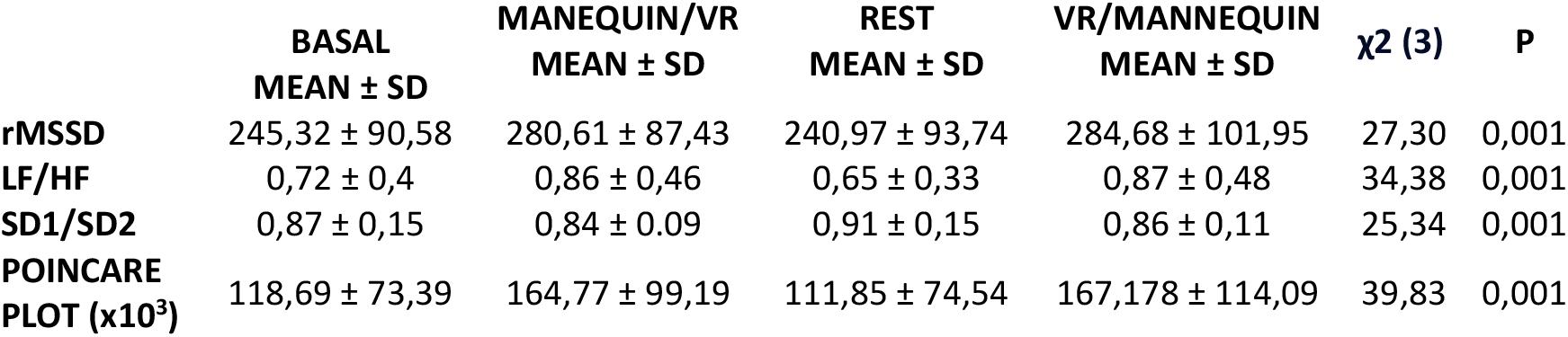
Biometric parameters at distinct stages of simulation (Friedman test)

**Fig. 1.**
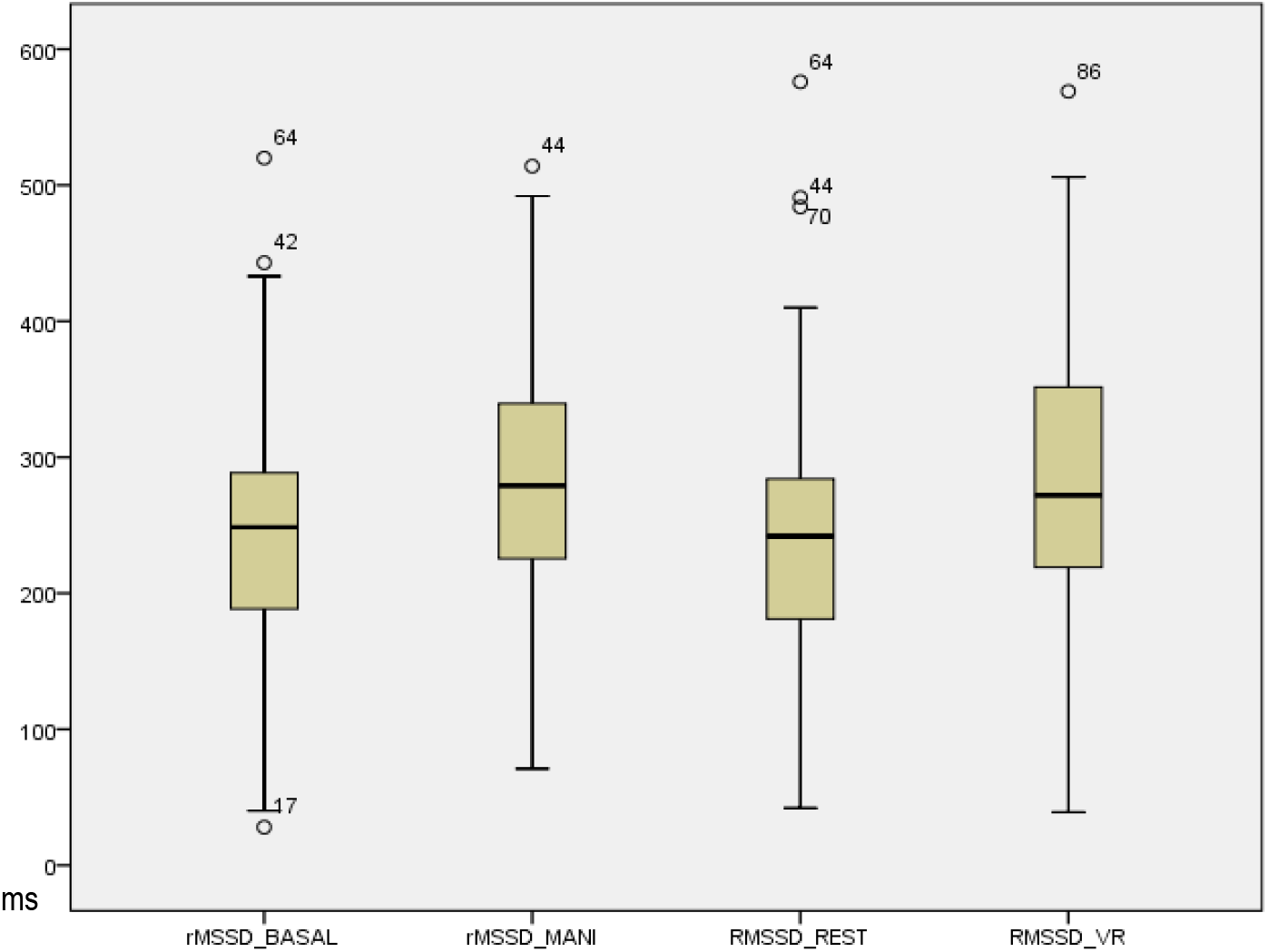
rMSDD values boxplot in the four scenarios (basal, mannequin, rest, and VR)

**Fig. 2.**
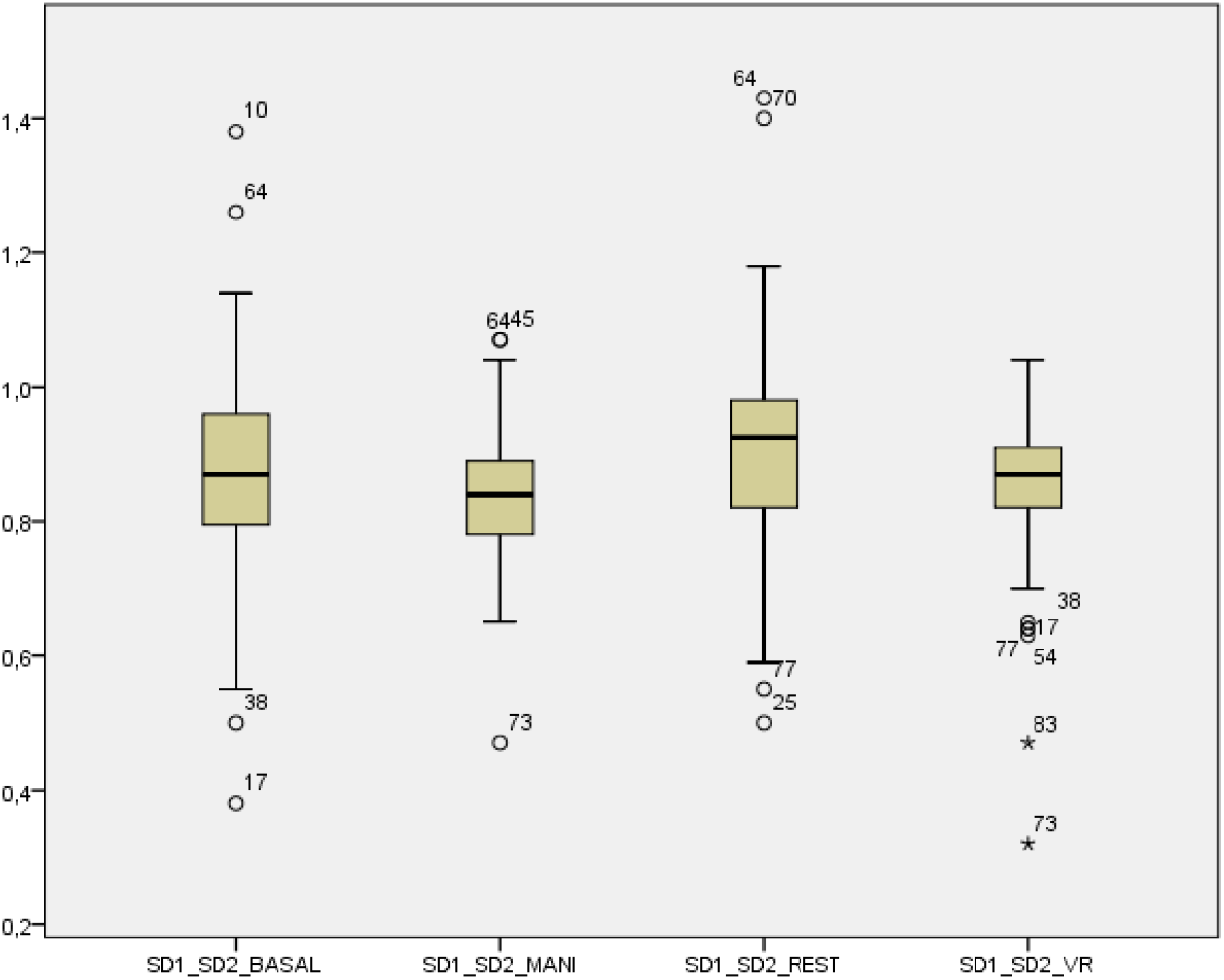
SD1/SD2 values boxplot in the four scenarios (basal, mannequin, rest, and VR)

Furthermore, we conducted a Wilcoxon signed-rank test to compare the HRV parameters between the two rest phases (basal and resting) and, more critically, between the VR and mannequin simulation phases. The analysis showed no significant differences between the stress responses elicited by the VR and mannequin simulations, indicating that both modalities equally effectively simulate the stress levels experienced in clinical scenarios. This equivalence in stress induction by both VR and traditional mannequin-based training highlights VR’s potential as a viable alternative in medical education, as documented in Table 5

**Table 5.** Biometric parameters paired (Wilcoxon signed-rank test)

|  | BASAL/REST (Z) | p | MANNEQUIN/VR(Z) | p |
| --- | --- | --- | --- | --- |
| rMSSD | -1,066 | 0,286 | -1,104 | 0,547 |
| LF/HF | -1,109 | 0,267 | -0,234 | 0,815 |
| SD1/SD2 | -3,065 | 0,002 | -1,941 | 0,52 |
| POINCAIRE PLOT | -2,026 | 0,43 | -0,4 | 0,968 |

### Bias and Sequence Effect Analysis

A detailed analysis was conducted to address potential selection biases and the effects of the sequence of simulation exposure. The data confirmed that the order of engagement—whether participants started with VR or with the mannequin—did not significantly impact stress outcomes. This robustness supports the validity of our experimental design and substantiates the equivalence of both training modalities in terms of their stress-inducing capabilities (Tables 6 and 7).

**Table 6.** Biometric parameters (First VR Session, Friedman test).

| | BASAL<br>MEAN ± SD | VR<br>MEAN ± SD | REST<br>MEAN ± SD | MANNEQUIN<br>MEAN ± SD | $\chi^2$ (3) | p |
| --- | --- | --- | --- | --- | --- | --- |
| rMSSD | 250,91 ± 84,73 | 290,41 ± 87,55 | 251,72 ± 87,75 | 301,59 ± 111,75 | 13,09 | 0,004 |
| LF/HF | 0,70 ± 0,32 | 0,83 ± 0,24 | 0,66 ± 0,36 | 0,91 ± 0,47 | 25,38 | 0,001 |
| SD1/SD2 | 0,87 ± 0,12 | 0,85 ± 0,1 | 0,92 ± 0,13 | 0,84 ± 0,12 | 14,20 | 0,003 |
| POINCARE<br>PLOT ($\times 10^3$) | 124,12 ± 75,59 | 170,59 ± 95,74 | 119,18 ± 70,55 | 190,74 ± 128,47 | 19,29 | 0,001 |

**Table 7.** Biometric (First Mannequin Session, Friedman Test)

| | BASAL<br>MEAN ± SD | MANNEQUIN<br>MEAN ± SD | REST<br>MEAN ± SD | VR<br>MEAN | STD DEV | $\chi^2$ (3) | p |
| --- | --- | --- | --- | --- | --- | --- | --- |
| rMSSD | 239,74 ± 98,55 | 270,81 ± 97,02 | 230,22 ± 99,02 | 267,76 | 88,96 | 16,31 | 0,001 |
| LF/HF | 0,73 ± 0,46 | 0,89 ± 0,61 | 0,65 ± 0,3 | 0,84 | 0,49 | 11,13 | 0,011 |
| SD1/SD2 | 0,87 ± 0,17 | 0,83 ± 0,08 | 0,90 ± 0,16 | 0,87 | 0,09 | 13,40 | 0,04 |
| POINCARE<br>PLOT ($\times 10^3$) | 113,26 ± 71,41 | 158,95 ± 103,12 | 104,52 ± 78,31 | 143618,67 | 92978,90 | 22,33 | 0,001 |

### Secondary Analysis of the Stress Parameters

This secondary analysis delved into the intricate relationships between stress responses and a range of demographic and lifestyle factors, employing repeated-measures ANOVA with Bonferroni and Tukey post hoc tests to rigorously examine these interactions.

#### Age and Gender Influences

We observed significant interactions between the HRV parameter RMSSD and the combined factors of age and gender (Table 8). This highlights the importance of demographic variations in assessing physiological stress responses in medical education, suggesting that different age groups and genders may experience stress differently during simulations.

**Table 8.** Multivariate analysis of variance results for RMSSD by age and sex.

| Variable | F | df1/df2 | $\eta^2$ | p |
| --- | --- | --- | --- | --- |
| RMSSD | 8,175 | 3,0/91 | 169422,779 | 0,001 |
| RMSSD * AGE | 1,648 | 30,0/267,78 | 242440,437 | 0,002 |
| RMSSD * SEX | 1,805 | 3,0/91,3 | 29816,49 | 0,64 |
| RMSSD * AGE * SEX | 2,566 | 9,00/221,62 | 95792,567 | 0,008 |

#### Lifestyle dynamics

Our findings indicated that lifestyle variables, particularly marital status, living conditions, and family income, significantly influenced stress responses. Notably, the interaction between family income and work-study balance was strongly correlated with the LF/HF ratio, a marker of sympathetic activity, suggesting that economic factors may exacerbate stress levels among medical students (Table 9).

**Table 9.**
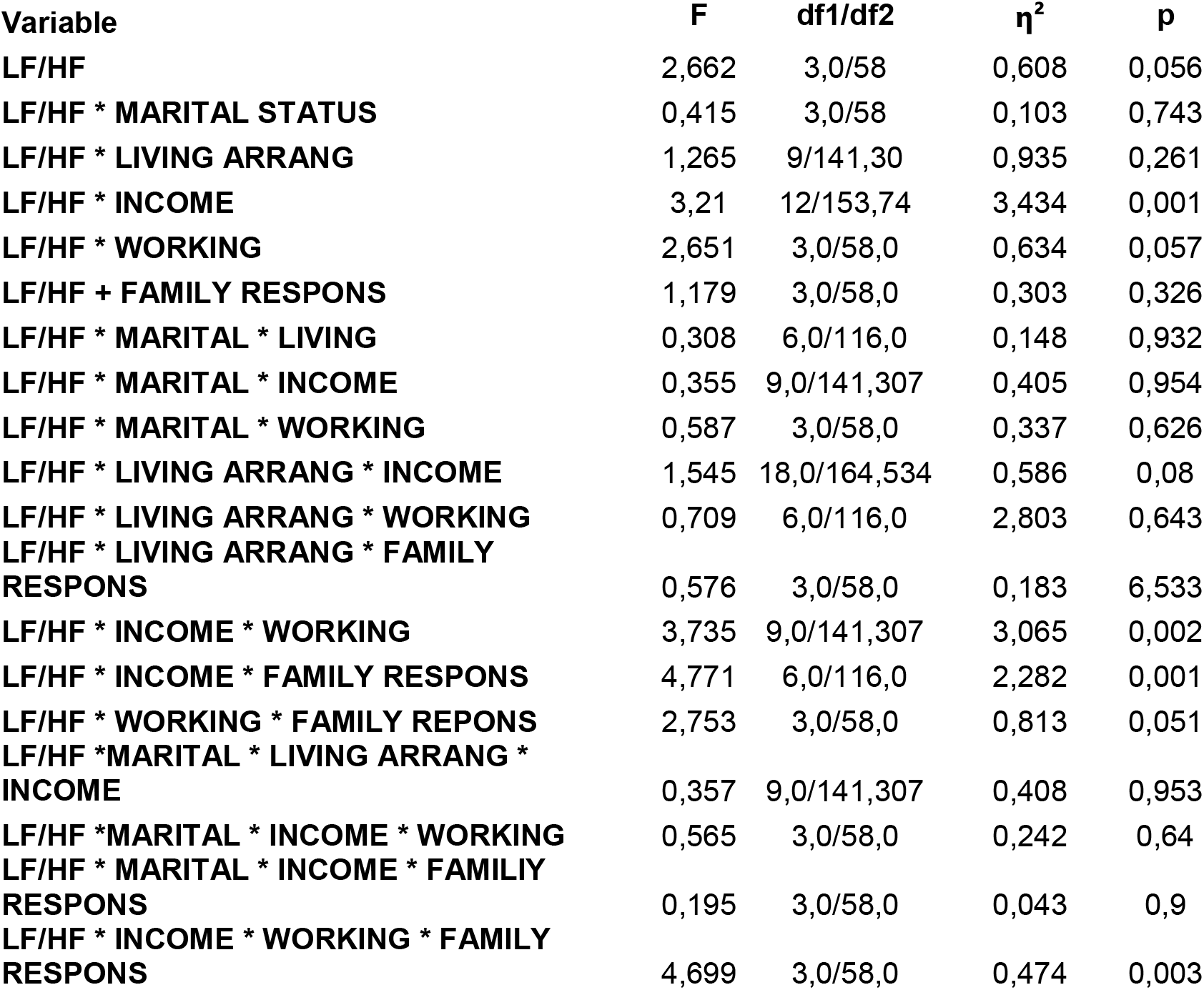
Multivariate analysis of variance results for LF/HF stratified by marital status, living arrangement, family income, working and studying and family responsibilities.

#### Consumption Habits

The impact of consumption habits such as smoking, alcohol use, and caffeine intake on stress was less pronounced, with minimal influence on primary stress markers. However, subtle interactions were noted, indicating potential areas for further investigation.

#### Medication Impact

The combined use of anxiolytics and antidepressants showed complex interactions that significantly affected stress biomarkers, underscoring the need to consider medication history when analyzing stress responses in medical training settings.

#### Academic and Perceived Stress

Contrary to initial expectations, our study revealed no significant correlations between study habits, perceived academic stress, and physiological stress markers among last-year medical students. This finding suggests that these students, having progressed through rigorous academic training over several years, may have developed effective coping mechanisms to manage and mitigate the physiological manifestations of stress. This adaptation could explain why academic demands and self-perceived stress do not markedly alter physiological stress responses in this group. However, variations might exist with different student populations or in distinct educational settings, indicating a potential area for further research to explore how stress coping mechanisms evolve throughout medical education.

## DISCUSSION

Our study suggests that virtual reality (VR) is comparable to traditional mannequin-based training in simulating stress responses during pericardiocentesis. While VR offers immersive and interactive environments, our findings cautiously support its equivalence rather than superiority over traditional methods. This aligns with research by Tsun-Ying and Marvin Mergen, who noted VR’s capability in procedural training without conclusively establishing its enhancement of clinical decision-making skills [3].

Consistent with Bolton’s findings, VR shows promise in enhancing access to medical training in low- and middle-income countries [9]. However, its broader application, including overcoming geographical and economic barriers, must be approached with a focus on sustainability and adaptability to local contexts. Concerns raised by Weissgalss regarding medical artificial intelligence underscore the need for robust policy frameworks to manage potential biases [10].

Our study also explored the integration of 3D modeling with VR, which enhances both the cost-effectiveness and educational accessibility of medical training. Ellen M. Hong and Tae Hoon Roh provided complementary evidence on the benefits of open-source software and photorealistic 3D models in surgical training [11] [12]. Such advancements support the broader application of these technologies in various medical disciplines, particularly in resource-limited environments.

Moreover, our research delves into the psychological aspects of medical training. By incorporating 3D-printed mannequins and VR, we address both the technical and emotional challenges faced by healthcare professionals. This dual-focus approach is corroborated by studies from Ashley Towers and Lauryn R [13]. Rochlen noted significant improvements in student confidence and procedural accuracy with VR and 3D technologies [14].

The application of VR in critical care settings, as discussed by Bruno, highlights its potential to enhance educational outcomes while also presenting challenges related to technology integration, cost, and ethical considerations [15]. These insights underscore the necessity of navigating technical and human factors to fully exploit the benefits of immersive technologies in high-stakes environments.

Our secondary analysis, which explored the relationships between stress parameters and demographic variables, revealed significant interactions between physiological biomarkers and age, sex, lifestyle, and medication intake. Tessa Helman’s work provides an important context for these findings, highlighting how stress responses can differ significantly between genders, potentially influencing the development of conditions such as coronary artery disease [16]. These insights emphasize the complexity of physiological responses to stress and underscore the importance of personalized training approaches that consider individual demographic and lifestyle factors.

Moreover, the impact of lifestyle dynamics on physiological stress markers highlighted in our study emphasizes the complex interplay between socioeconomic factors and health. The findings suggest that family income and work-study balance significantly influence stress-related physiological responses, indicating the need for comprehensive lifestyle assessments in medical training. This finding aligns with Reinhold Kreutz’s observations on the effects of lifestyle changes induced by the COVID-19 pandemic on blood pressure and hypertension, further illustrating the interconnectedness of lifestyle factors and health outcomes [17].

Finally, our inclusion of heart rate variability (HRV) analysis in VR simulations provides a novel approach for assessing stress responses, which is critical for preparing medical professionals to perform under pressure. This aspect of our study aligns with findings from Sean L. Corrigan and David Narciso [18], who emphasized VR’s ability to replicate and manage real-world stress conditions effectively [19].

This study contributes to the understanding of VR’s potential in medical education, particularly for complex procedures. It emphasizes the need for careful integration of new technologies in training programs and highlights the importance of addressing infrastructural and ethical challenges to maximize their impact.

This study’s limitations are notable and must be carefully considered when interpreting the findings. First, the nonrandomized design may introduce biases that could affect the robustness of the conclusions. The context of a high-income country also limits the generalizability of our results to settings with different economic conditions. Importantly, our participant group consisted exclusively of undergraduate medical students, which raises concerns about the applicability of our findings to practicing healthcare professionals. The experiences and stress responses of students may differ significantly from those of seasoned professionals who have developed more advanced coping mechanisms and clinical skills. Furthermore, focusing solely on a single training model restricts our understanding of how various VR platforms and technologies might perform under different educational scenarios

Conversely, the study’s strengths are notable. The combination of traditional simulation with VR technologies fills a critical gap in medical education, making high-fidelity clinical simulation more accessible and reducing costs. Clinical validation by experienced physicians enhances the reliability of our findings and their practicality in real-world settings. The large sample size and objective methods used for assessing learning and stress parameters further solidify the study’s conclusions.

Future research should aim to expand the geographic and demographic scope of studies, exploring VR’s utility and effectiveness across a broader spectrum of medical training environments. Randomized controlled trials and longitudinal studies are needed to provide more definitive evidence of VR’s impacts on learning outcomes and clinical performance.

## CONCLUSION

This study highlights the effectiveness of virtual reality in simulating the stress-related aspects of complex medical procedures, such as pericardiocentesis. It demonstrates VR’s capability to replicate both the technical challenges and emotional pressures found in real-life medical scenarios, thereby significantly contributing to stress management training in medical education. VR’s immersive environments are instrumental in preparing medical students for the psychological demands they will face in their careers.

The secondary analysis reveals that demographic and lifestyle factors significantly influence stress responses during training, with VR’s adaptability allowing for personalized scenarios to effectively address these varied responses. However, the study’s limitations, including its nonrandomized design and focus on undergraduate students, necessitate cautious interpretation of the findings, and suggest limited generalizability to practicing healthcare professionals.

In conclusion, VR is evolving as a crucial tool in medical education, especially for stress management. Future research should aim to validate VR’s effectiveness in diverse and realistic settings and explore its potential across different medical training stages. Further studies are needed to integrate comprehensive stress analysis within VR training, tailoring it to the specific needs of medical trainees worldwide

## Data Availability

The datasets used and/or analyzed during the current study are available in publicly available repositories.

## DECLARATIONS

### Ethics approval and consent to participate

This study received ethical approval from the HM Montepríncipe University Hospital Research Ethics Committee (Code: 18.12.1339.GHM) and involved 132 consenting medical students. All participants provided written informed consent, and their data were anonymized to protect confidentiality

### Consent for publication

Not applicable

### Authors’ contributions

ARL designed the study, collected the data, performed the statistical analysis, and wrote the study as the principal investigator.

RG participated in the development and correction of the virtual scenario.

LZ performed the programming of the virtual scenario according to the created model.

ARN participated in the performance tests and in the design and collection of biometric parameters

AGP participated in the design and adaptation of the simulated scenarios

PCF participated in proofreading and supervision of the study design, statistical analysis and writing of the paper.

## Acknowledgments

Not applicable

